# Decline in striatal binding ratio associated with accelerated decline in performance on Symbol Digit Modality but not MoCA in Parkinson’s disease psychosis

**DOI:** 10.1101/2024.08.01.24311353

**Authors:** Sara Pisani, Latha Velayudhan, Dag Aarsland, K. Ray Chaudhuri, Clive Ballard, Dominic ffytche, Sagnik Bhattacharyya

## Abstract

**Background:** Cognitive deficits have been reported in Parkinson’s Disease psychosis (PDP). Reduced dopamine transporter (DAT) binding ratio has also been associated with PDP. However, it remains unclear whether DAT striatal binding ratio (SBR) may contribute to worsening cognitive performance in PDP. Here, we examined this using data from the Parkinson’s Progression Markers Initiative study.

**Methods:** We analysed data from 408 PD patients, from baseline to year 4 follow up, and classified patients into PD with (PDP) and without psychosis (PDnP). DAT SBR was available from DaTSCAN imaging with ^123^I-FP-CIT-SPECT. We examined all cognitive measures assessed at each time point, socio-demographics, neuropsychiatric and PD-specific symptoms were entered as covariates of interest.

**Results:** PDP patients had lower DAT SBR compared to PDnP patients (b=-0.092, *p*=0.035) which remained significant after controlling for age, sex, and ethnicity. PDP patients also reported worse trajectory of task performance on MoCA (b=-0.238, *p*=0.001) and Symbol Digit Modality (b=-0.534, *p*=0.016) across four years compared to PDnP patients. Worsening of MoCA scores in PDP was independent of DAT SBR decline (interaction group * study years, b=-0.284, *p*=0.016; three-way interaction group*study years*DAT SBR, b=0.127, *p*=0.225). However, declining performance in Symbol Digit Modality was significantly associated with the decline in DAT SBR (three-way interaction group*study years*DAT SBR, b=0.683, *p*=0.028).

**Conclusion:** Overall, longitudinal decline in striatal presynaptic dopamine function may underlie the greater longitudinal decline in performance in the symbol digit modality task that engages processing speed, associative learning and working memory in PD psychosis, whilst declining performance on MoCA seems unrelated to it. Whether striatal presynaptic dopamine changes explain accelerated longitudinal decline in other cognitive domains in people with PDP remains to be tested.

## Introduction

Psychotic symptoms such as hallucinations and delusions are common non-motor manifestations of Parkinson’s disease (PD) and are associated with increased burden of care, poor quality of life and risk of dementia ^1–4^. Consistent with the characteristic degeneration of nigrostriatal dopaminergic neurons in PD, lower striatal dopamine transporter (DAT) availability, a proxy measure of presynaptic dopamine deficiency ^5–7^, and indexed by striatal DAT binding ratio (a ratio of the specific binding concentration in the striatum to non-specific DAT binding concentration in the reference brain region), has been shown to be associated with psychosis ^8–10^ and other neuropsychiatric symptoms ^11–13^ in PD. Evidence from longitudinal observation of newly diagnosed, de novo PD patients as part of the Parkinson’s Progression Markers Initiative (PPMI) ^14^, indicates that striatal DAT binding ratio declines over time in PD patients ^15^ ^16^. However, whether such a longitudinal decline is also associated with the occurrence of psychosis in PD and is similar or different from those who do not develop psychosis remains unclear ^1^ ^17^. A recent analysis of data from the PPMI cohort suggests an inverse relationship between apathy/anhedonia but not depression or motor symptoms in people with PD ^12^.

Consistent with the well-recognised role of dopamine in a range of cognitive processes including cognitive control, attention, cognitive flexibility, reasoning, language, and learning ^18–20^ and emerging evidence linking decline in presynaptic striatal dopamine function with age-related decline in cognitive performance ^21^ ^22^, low baseline striatal binding ratio has been associated with cognitive impairment (as indexed using Mini Mental State Examine (MMSE), and Montreal Cognitive Assessment (MoCA)) ^10^ in people with PD, and lower caudate and putamen binding ratio was associated with poor performance in frontal and executive (e.g., Frontal Assessment Battery, Trial Making Test), and visuo-spatial domains of cognition in people with PD and mild cognitive impairment (MCI) ^23^. Further, compared to those without MCI, PD patients with MCI have been shown to have lower caudate dopamine uptake that directly correlated with caudate functional activity ^24^. Independent evidence also suggests a positive correlation between striatal DAT binding and processing speed (measured with the Symbol Digit Modality, SDM) ^25^ and attention/executive function ^26^ and an inverse relationship with cognition-related metabolic pattern ^27^ in people with PD. A recent work by Qamar et al. ^28^ focused on systematically reviewing the evidence regarding the involvement of dopaminergic mechanisms across different non-motor symptoms. Their findings outlined reduced dopaminergic uptake in striatal regions, and in pre-frontal regions (known to be associated with executive functions) in PD-MCI and PDD patients compared to healthy controls. Similarly, PD patients with hallucinations had more dopaminergic depletion, expressed as lower DAT binding ratio, in the striatum compared to PD without perception disorders. While cross-sectional evidence summarised here may suggest that striatal (including caudate) dopamine depletion may be associated with some cognitive deficits in PD patients, how this relationship evolves over time as PD progresses and some people go on to develop psychotic symptoms, remains unclear. Cognitive impairments in people with PD psychosis have been extensively reported ^29^ ^30^. Consistent with clinical impression, emerging evidence suggests that PD patients who go on to develop psychosis may experience a greater longitudinal decline in cognition ^31^ ^32^. This is further supported by our recent analysis of data from the Parkinson’s Progression Markers Initiative (PPMI) cohort ^14^, wherein we have reported accelerated decline in semantic aspects of language, processing speed, general cognitive abilities, visuo-spatial ability, immediate and delayed recall in PD patients with psychosis (PDP) compared to those without psychosis (PDnP) that was unlikely to be a result of differential trajectory of depression, sleepiness, REM sleep behaviour disorder and severity of motor symptoms ^33^. However, although longitudinal decline in cognition is evident in PD psychosis patients, whether this is related to longitudinal changes in presynaptic dopamine availability remains to be determined. Therefore, we examined this using the data from the longitudinal cohort of the PPMI study^14^. Investigating a potential association between dopaminergic depletion and deficits in specific cognitive domains could further our understanding of the neurobiological mechanisms involved in PD psychosis and its cognitive dysfunctions. Based on evidence of low baseline striatal DAT binding ratio (hereafter referred to as SBR) in PD psychosis patients ^8–10^, we predicted a greater longitudinal decline of SBR in PD psychosis compared to that in PD patients without psychosis. As the sample analysed here is a subset of the larger PPMI sample (please see ‘Methods’ for more details), we then tested which of the cognitive domains showing an accelerated decline in PDP compared to PDnP in our previous analysis of the larger PPMI cohort ^33^ would continue to show a similar differential decline in the smaller sized dataset analysed here. We also predicted that for the cognitive domains showing greater decline in PDP compared to PDnP, the longitudinal trajectory of task performance will show a differential association with the longitudinal trajectory of DAT SBR based on condition (PDP vs PDnP).

## Methods

### Participants

The PPMI study enrolled newly diagnosed unmedicated patients with Parkinson’s Disease and age- and gender-matched healthy controls. Details of eligibility criteria, objectives and methodology have been published elsewhere ^14^ and can be also found on www.ppmi-info.org/study-design (ClinicalTrials.gov NCT01141023). Description of the patients and healthy controls involved in this analysis are reported in Marek et al. ^14^. These data were accessed and downloaded in January 2023; the data used in this analysis are openly available from the PPMI study. The PPMI study was approved by the institutional review boards at each site ^14^ ^34^, and the participants provided written informed consent. In brief, PD patients were included if they were drug-naïve and within 2 years of PD diagnosis, with a Hoehn and Yahr stage <3, if they were 30 years of age or older and had either at least two of resting tremors, bradykinesia or rigidity (must have either resting tremor or bradykinesia) or a single asymmetric resting tremor or asymmetric bradykinesia. Age-matched and sex-matched health controls were included if they were 30 years or older, and with no evidence of any neurological disorder or a first-degree relative with PD. Although, the size of the total PPMI cohort is n= 676, in order to address the specific objectives of the present study, we focused on all participants (n= 420) with complete data on the cognitive domains of interest (please see below) as well as on striatal DAT binding ratio. Please note that the sample included in the present analyses is smaller than the sample analysed in our study examining the trajectory of cognition in people with PDP ^33^.

### Outcome measures

Outcomes of interest included the longitudinal trajectory of DAT striatal binding ratio (SBR) and its relationship with the trajectory of cognitive performance in people with PD. We included the different cognitive tasks assessed in the PPMI cohort: the Montreal Cognitive Assessment (MoCA) ^35^, the Hopkins Verbal Learning Test – Revised (HVLT-R) ^36^, the Symbol Digit Modality (SDM) ^37^ ^38^, Letter Number Sequence (LNS) ^39^, Semantic Fluency tests ^40^, Benton Judgement of Line Orientation (BJLOT) ^41^; depressive symptoms assessed using the Geriatric Depression Scale 15 items (GDS-15) ^42^; anxiety assessed using the State-Trait Anxiety Inventory (STAI) ^43^ state subscale; sleep using the Epworth Sleepiness Scale; and sleep behaviour using a REM sleep behaviour disorder (RBD) questionnaire ^44 45^. PD severity was assessed using the Movement Disorders Society Unified Parkinson’s Disease Rating Scale (MDS-UPDRS), which includes measures of tremors and rigidity, and Hoehn & Yahr stages ^46^. PD medications were expressed as Levodopa equivalent daily dose (LEDD). Information about LEDD was extracted from the most recently collected data and followed the recommendation of the PPMI study group (see Supplementary Material 1 eTable1 for more information, and for group differences on LEDD). Autonomic symptoms were measured with the Scale for Outcome in Parkinson’s Disease – Autonomic (SCOPA-AUT) ^47^.

### Classification of PD psychosis

PD patients were classified into PDP and PDnP based on previous work ^48^. In brief, we applied the MDS-UPDRS part I hallucinations/psychosis item, which measures the presence of visual hallucinations and paranoid thoughts. If PD patients reported a score ≥1 on the UPDRS part I hallucination/psychosis item at any study visits, they were considered as PDP (i.e., PD with psychosis). We have excluded PD patients who reported psychosis symptoms at baseline (n=12) as we wanted both groups to start off on a similar level (see Supplementary Material 1 eTable2-4 for more information). Therefore, three groups were therefore created:

- *HC:* healthy controls (no PD symptoms)
- *PDnP:* PD patients with a score of 0, i.e., without reports of psychotic symptoms throughout the study
- *PDP:* PD patients with a score ≥1, i.e., with presence of minor psychotic symptoms (illusions) and more moderate or severe symptoms (hallucinations or delusions), at any time point from year 1 follow up (inclusive) throughout the study.

### Image processing and calculation of striatal DAT binding ratio

The images used in this analysis were derived from ^123^I-FP-CIT-SPECT imaging at all time points from baseline (year 0) to year 4 follow up. Images were analysed according to the PPMI imaging protocol (details can be found at https://www.ppmi-info.org/study-design/research-documents-and-sops), and, as region of interest (ROI), we focused on bilateral striatum. ROI included bilateral caudate and bilateral putamen which provided the summative measure of bilateral striatum. Count densities were obtained and used in computing the striatal binding ratio. DAT SBR was determined by the formula (target region/reference region)-1, with occipital region as reference region. We have not included healthy controls in this study as DAT SBR was available for this group only at baseline, whilst DAT SBR was available for PD patients for all four time points after baseline.

### Statistical analysis

Baseline characteristics of the cohort included in the present study were compared using independent t test or its non-parametric equivalent (e.g., Mann-U Whitney test) as appropriate. Bonferroni correction for multiple comparisons was applied. We report baseline summaries for socio-demographic characteristic, HVLT-R, SDM, semantic fluency test, LNS, BJLOT, MoCA, neuropsychiatric measures (such as depression and RBD), and PD-related assessments. We examined all study visits from baseline to follow-up year 4. However, for the purpose of this analysis, we only focused on PDP and PDnP patients who underwent ^123^I-FP-CIT-SPECT imaging at each time point during their participation in the PPMI study, as healthy control participants underwent such examination only at baseline. We employed linear mixed-effect model with restricted maximum likelihood (REML) using the *lmerTest* package in R (version 4.0.3) ^49^ with group as between-subject factor, year as within-subject predictor which was considered as a continuous measure for the present analyses, and patient number as random factor to examine the longitudinal trajectory of presynaptic dopamine levels in the striatum as indexed by SBR. Significance (a two-sided alpha level of 0.05) was estimated using Satterthwaite’s method. We first tested an unadjusted model and then an adjusted model where we included the following covariates of interest: sociodemographic characteristics; scores on depression, sleepiness, RBD, motor symptoms; and a three-way interaction term including PD groups, study years and SBR. We included scores at each time point for all variables and covariates of interest. PD medications expressed in LEDD (mg/day) were not included in the model, as we did not find any differences in the amount of LEDD or in the trajectory of dopamine-replacement medications between PDnP and PDP patients (please refer to Supplementary Material 1, eTable1). The adjusted analysis including the three-way interaction (group * study year * DAT SBR) was conducted only on the cognitive domains for which we observed a significantly different longitudinal trajectory between PDP and PDnP patients in the sample analysed here (n=420).

## Results

### Baseline sample characteristics

Table 1 reports baseline characteristics of PDP and PDnP patients. There were no significant differences across age, sex, years of education, duration, and age of onset of PD and age of PD diagnosis between the two groups. There were more patients identifying as “white” in both groups, compared to other ethnic groups (*p*=0.021). PDP patients reported more depressive symptoms (*p*=0.035), more RBD (*p*=0.019), more severe symptoms as reported with MDS-UPDRS part 1 (*p=*0.001) and part 2 (*p*=0.012), and more severe autonomic symptoms (*p*<0.001) compared with PDnP patients. There were no differences between groups on any cognitive measures. For group differences at baseline between PD patients and those who developed psychotic symptoms at baseline, please refer to Supplementary Material 1, eTable2-4.

**Table 1.**
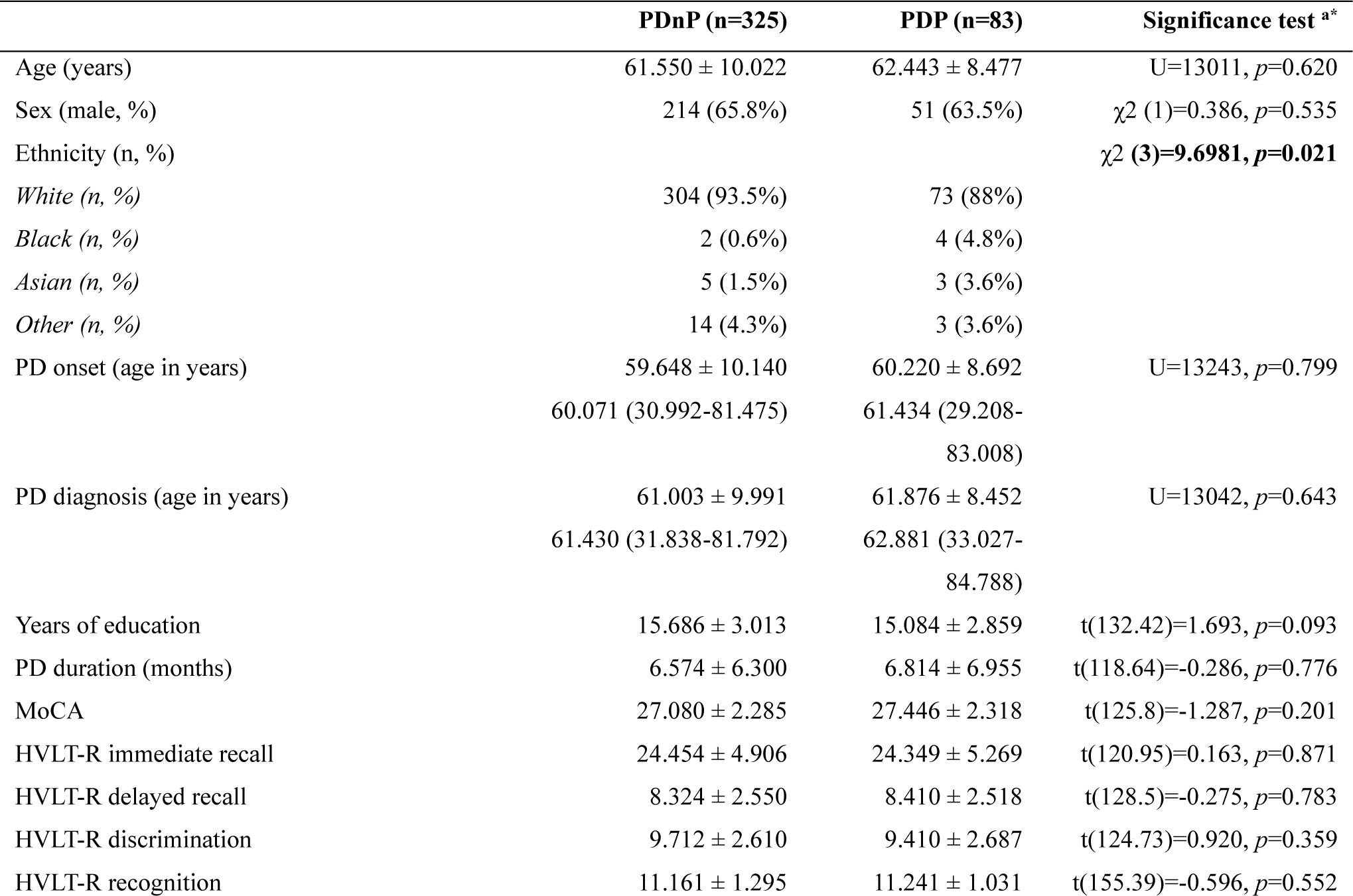
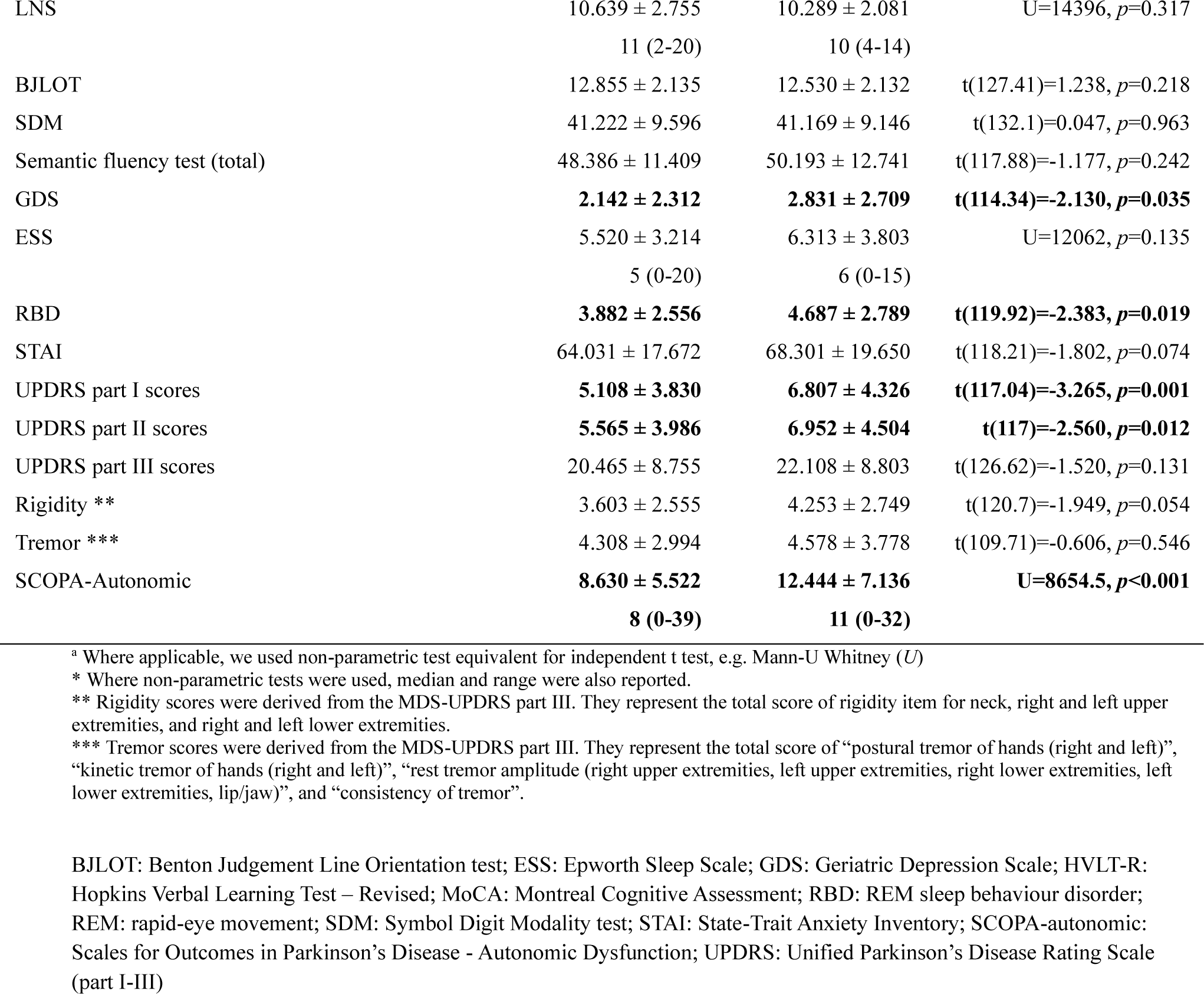
Sample baseline characteristics of the two groups, i.e., PD patients without psychosis (PDnP), and PD patients with psychosis (PDP). PDP psychosis group does not include PD patients who reported psychosis symptoms at baseline (please refer to Supplementary Material 1 eTable4). Means and standard deviations are reported, unless specified otherwise.

### Trajectory of striatal DAT binding ratio in PD psychosis

Unadjusted analysis showed a significant main effect of group with PDP patients having reduced DAT SBR compared with PDnP (b=-0.092, 95% CI, -0.183, -0.007, *p*=0.035) across all of the follow-up time-points. There was also a main effect of time (i.e., study year) showing decline of DAT striatal binding ratio across the 4 years in both groups (b=-0.094, 95% CI, -0.101, -0.087, *p*<0.001), however there was no significant interaction group * time (b=-0.009, 95% CI, -0.023, 0.005, *p*=0.214). After controlling for age, sex and ethnicity, results did not change (main effect of group, *p*=0.040; main effect of time, *p*<0.001; interaction, *p*=0.214) (Figure 1).

**Figure 1.**
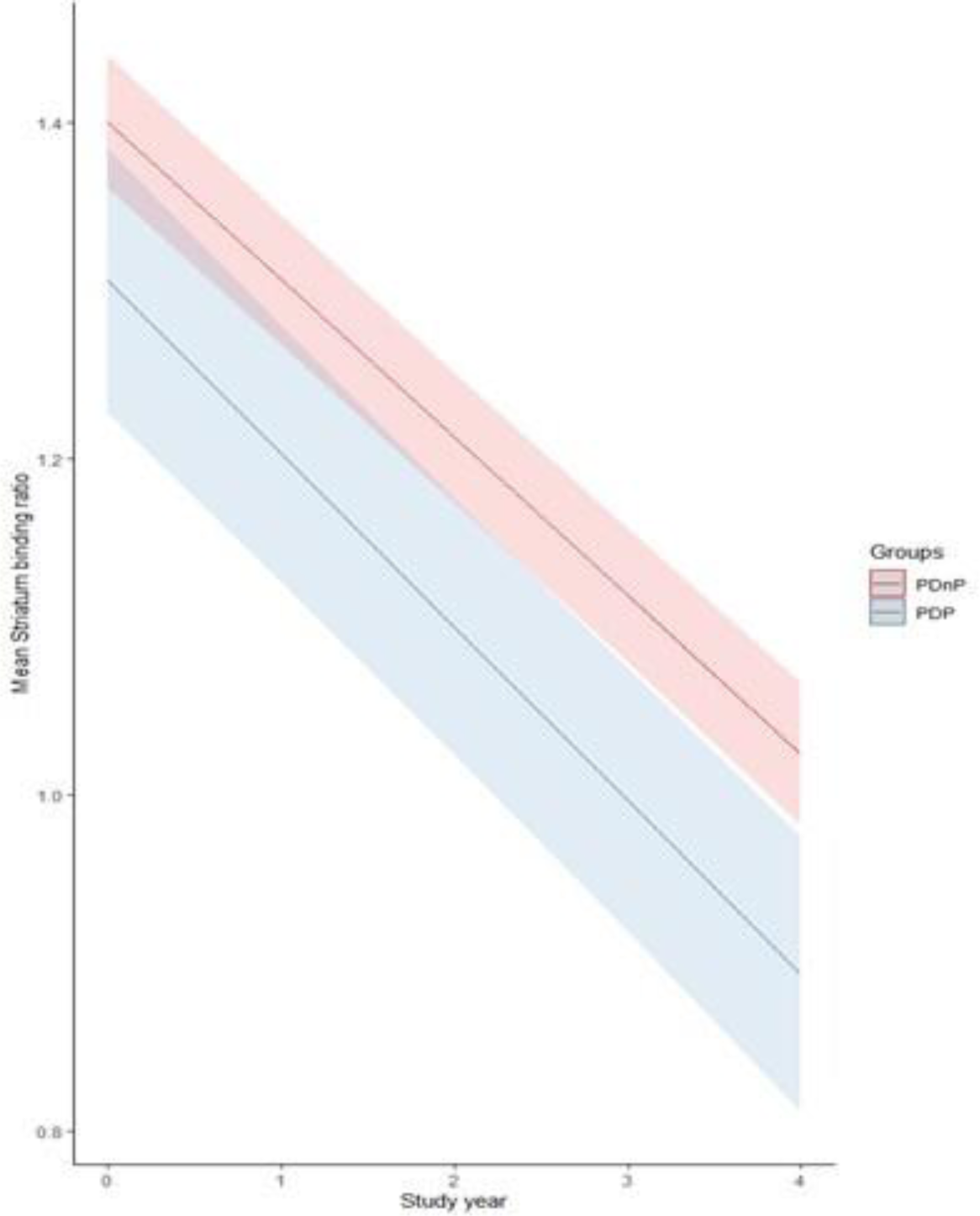
Trajectory of DAT striatal binding ratio in PD patients with (PDP) and without psychosis (PDnP) in bilateral striatum across the 4-year of the PPMI study.

### Trajectory of cognitive performance in PD psychosis

We have already reported results from our study examining the longitudinal trajectory of cognition in PDP ^33^ in the larger cohort of PPMI patients. As the present investigation primarily focuses on the differential relationship between the longitudinal trajectory of presynaptic dopamine (DAT SBR) and the longitudinal trajectory of cognition in PDP versus PDnP patients, for the sake of completeness we also report here the longitudinal trajectory cognition in the subset of PPMI patients with DAT SBR who are the focus of the present investigation.

Unadjusted analyses revealed that, across most of the cognitive tests, there were no differences or differential longitudinal trajectories between PDP and PDnP patients. However, there was a significant group*time interaction for performance on the MoCA (b=-0.238, 95% CI, -0.375, -0.100, *p*=0.001), whereby PDP patients showed a worse trajectory in performance compared with PDnP patients. This indicates that for each additional year of follow up from baseline, the MoCA total score in PDP patient is 0.187 points less than that of PDnP patients. Similarly, there was a significant interaction group*time interaction in SDM (b=-0.534, 95% CI, -0.969, -0.100, *p*=0.016) whereby PDP patients reported a worse trajectory in SDM performance over 4 years compared with PDnP patients. No other interactions or main effects were significant (Table 2). Please note that these results are different from our study examining the longitudinal trajectory of cognition in PDP ^33^, which examined the larger cohort of patients and showed an accelerated decline across the domains of visuo-spatial abilities (measured with BJLOT), semantics aspects of language (as indexed by semantic fluency performance), immediate and delayed recall (assessed with the HVLT-R), in addition to MoCA and SDM (reported in the present study) in PDP compared to PDnP patients.

**Table 2.** Main effect of group for PDnP and PDP, and interaction group * time for from the unadjusted analysis are reported here for all cognitive measures. Regression coefficient (estimate, b) with 95% confidence intervals (95% CI) and associated *p* value are reported. Highlighted in grey are results that are statistically significant pertaining to the main effect and interaction (group * time).

|  | Main effect of group (PDP vs PDnP) |  | Main effect of time (study year) |  | Interaction (group * time) † |  |
| --- | --- | --- | --- | --- | --- | --- |
|  | Estimate (95% CI) | *p* value | Estimate (95% CI) | *p* value | Estimate (95% CI) | *p* value |
| HVLT-R Immediate Recall | -0.162 (-1.380, 1.056) | 0.795 | 0.021 (-0.101, 0.143) | 0.739 | -0.162 (-0.421, 0.97) | 0.220 |
| HVLT-R Delayed Recall | 0.187 (-0.452, 0.825) | 0.566 | -0.007 (-0.072, 0.057) | 0.820 | -0.120 (-0.264, 0.010) | 0.069 |
| HVLT-R Recognition | -0.007 (-0.286, 0.272) | 0.963 | -0.004 (-0.049, 0.041) | 0.863 | 0.011 (-0.085, 0.107) | 0.826 |
| HVLT-R Discrimination | -0.107 (-0.595, 0.381) | 0.667 | **0.174 (0.101, 0.246)** | **<0.001** | 0.054 (-0.101, 0.208) | 0.495 |
| BJLOT | -0.297 (-0.794, 0.200) | 0.241 | **0.003 (-0.0511, 0.057)** | **0.905** | 0.055 (-0.060, 0.170) | 0.349 |
| Semantic Fluency | 1.269 (-1.478, 4.016) | 0.365 | -0.108 (-0.323, 0.106) | 0.322 | -0.177 (-0.632, 0.278) | 0.445 |
| **SDM** | -0.350 (-2.749, 2.049) | 0.775 | **-0.378 (-0.584, -0.174)** | **<0.001** | **-0.534 (-0.969, -0.100)** | **0.016** |
| LNS | -0.243 (-0.862, 0.376) | 0.442 | **-0.089 (-0.149, -0.030)** | **0.003** | -0.026 (-0.153, 0.101) | 0.690 |
| **MoCA** | 0.320(-0.304, 0.945) | 0.315 | -0.056 (-0.121, 0.008) | 0.088 | **-0.238 (-0.375, -0.100)** | **0.001** |

†Interaction ‘group \* time’ refers to the interaction PDP \* Study years with PDnP as the reference group in our comparisons.
BJLOT: Benton Judgement Line Orientation test; HVLT-R: Hopkins Verbal Learning Test – Revised; LNS: Letter Number Sequence; MoCA: Montreal Cognitive Assessment; SDM: Symbol Digit Modality test

### Association between longitudinal decline in DAT SBR and longitudinal decline in MoCA and Symbol Digit Modality performance in PD psychosis

In the present sample, as significantly worse performance over time in PDP compared with PDnP was evident for MoCA and SDM, we conducted two three-way interaction (group * study year * DAT SBR) analyses, one with MoCA and another with SDM as outcome measures. These were also adjusted for socio-demographics at baseline, as well as depression, sleep-related issues, and motor symptom severity scores over all the time-points. Results of the three-way mixed-effect models with MoCA and one with SDM as outcome measures are summarised in Table 3.

**Table 3.** Adjusted analysis controlling for depression, sleepiness, REM sleep behaviour disorder, and motor symptoms (i.e., UPDRS part 3). Regression coefficient (estimate, b) with 95% confidence intervals (95% CI) and associated *p* value are reported for all analyses. Highlighted in grey are results that are statistically significant pertaining to the main effect and interaction (group * time, group * time * DAT SBR).

|  | MoCA |  |  | SDM |  |  |
| --- | --- | --- | --- | --- | --- | --- |
|  | Estimate | 95% CI | *p* value | Estimate | 95% CI | *p* value |
| **Main effect of group, time (year), striatal binding ratio** |  |  |  |  |  |  |
| PDP vs PDnP | 0.568 | -0.010, 1.146 | 0.054 | 1.356 | -0.673, 3.386 | 0.190 |
| Year | 0.044 | -0.065, 0.152 | 0.432 | 0.023 | -0.313, 0.360 | 0.892 |
| Striatal binding ratio | 0.204 | -0.023, 0.432 | 0.078 | 0.392 | -0.347, 1.130 | 0.298 |
| Age | **-0.771** | **-0.981, -0.561** | **<0.001** | **-4.332** | **-5.091, -3.572** | **<0.001** |
| Sex | **0.353** | **0.147, 0.559** | **0.001** | **1.554** | **0.806, 2.302** | **<0.001** |
| Ethnicity | **-0.457** | **-0.655, -0.260** | **<0.001** | -0.627 | -1.348, 0.094 | 0.088 |
| Education (years) | **0.333** | **0.131, 0.535** | **0.001** | **1.913** | **1.181, 2.644** | **<0.001** |
| GDS | -0.064 | -0.239, 0.111 | 0.472 | **-0.891** | **-1.447, -0.335** | **0.002** |
| ESS | -0.147 | -0.315, 0.022 | 0.088 | -0.457 | -0.990, 0.076 | 0.093 |
| RBD | 0.006 | -0.161, 0.174 | 0.942 | **-0.537** | **-1.070, -0.004** | **0.048** |
| UPDRS part 3 | **-0.275** | **-0.445, -0.105** | **0.002** | **-1.194** | **-1.730, -0.658** | **<0.001** |
| **Interactions (group\*time\*DAT striatal binding ratio)** |  |  |  |  |  |  |
| PDP * PDnP (year) | **-0.284** | **-0.514, -0.053** | **0.016** | -0.186 | -0.897, 0.524 | 0.607 |
| PDP * PDnP (Striatal binding ratio) | -0.456 | -0.984, 0.073 | 0.091 | -0.468 | -2.192, 1.256 | 0.594 |
| YEAR * Striatal binding ratio | -0.082 | -0.182, 0.019 | 0.112 | -0.250 | -0.552, 0.052 | 0.104 |
| PDP * YEAR * Striatal binding ratio | 0.127 | -0.078, 0.331 | 0.225 | **0.683** | **0.073, 1.293** | **0.028** |

ESS: Epworth Sleep Scale; GDS: Geriatric Depression Scale; MoCA: Montreal Cognitive Assessment; RBD: REM sleep behaviour disorder; REM: rapid-eye movement; PDnP: PD patients without psychosis; PDP: PD patients with psychosis; SDM: Symbol Digit Modality test; UPDRS: Unified Parkinson's Disease Rating Scale (part 3)

### MoCA

We found a significant interaction between group * time (b=-0.284, 95% CI, -0.514, - 0.053, *p*=0.016) but no significant three-way interaction (i.e., group * time * striatum, *p*=0.225) indicating an accelerated longitudinal decline in MoCA performance in PDP compared with PDnP that remained significant even after controlling for the differential longitudinal trajectories of SBR in the two groups (entered as an interaction term: group * time * SBR) as well as potential confounding effects of baseline sociodemographic characteristics and longitudinal course of depression, sleep changes, and motor symptom severity (Figure 2).

**Figure 2.**
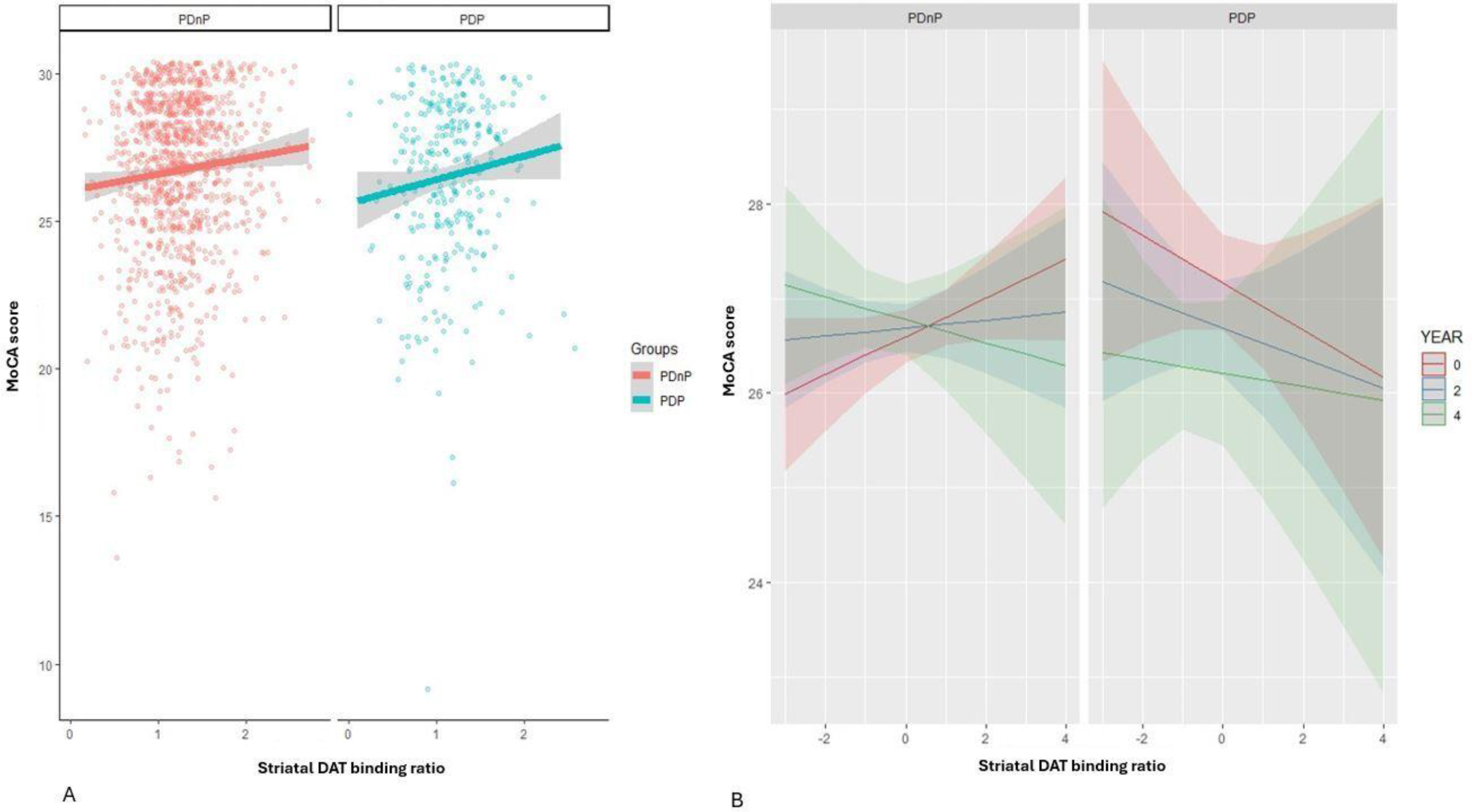
Trajectory of performance in the MoCA in PD patients with (PDP) and without psychosis (PDnP) over the years of the PPMI study and DAT SBR (A), and predictive values of MoCA for each PD group (B).

### Symbol Digit Modality

There was a significant three-way interaction between group * time * SBR indicating that the accelerated longitudinal decline in SDM performance in PDP patients compared to PDnP patients was associated with the differential longitudinal trajectories of decline in striatal binding ratio (SBR) in the two groups even after controlling for potential confounding effects of baseline sociodemographic characteristics and longitudinal course of depression, sleep changes, and motor symptom severity (b=0.683, 95% CI, 0.073, 1.293, *p*=0.028). Figure 3 shows the relationship between striatal DAT binding ratio with SDM performance in PDP and PDnP patients (across the 4 years).

**Figure 3.**
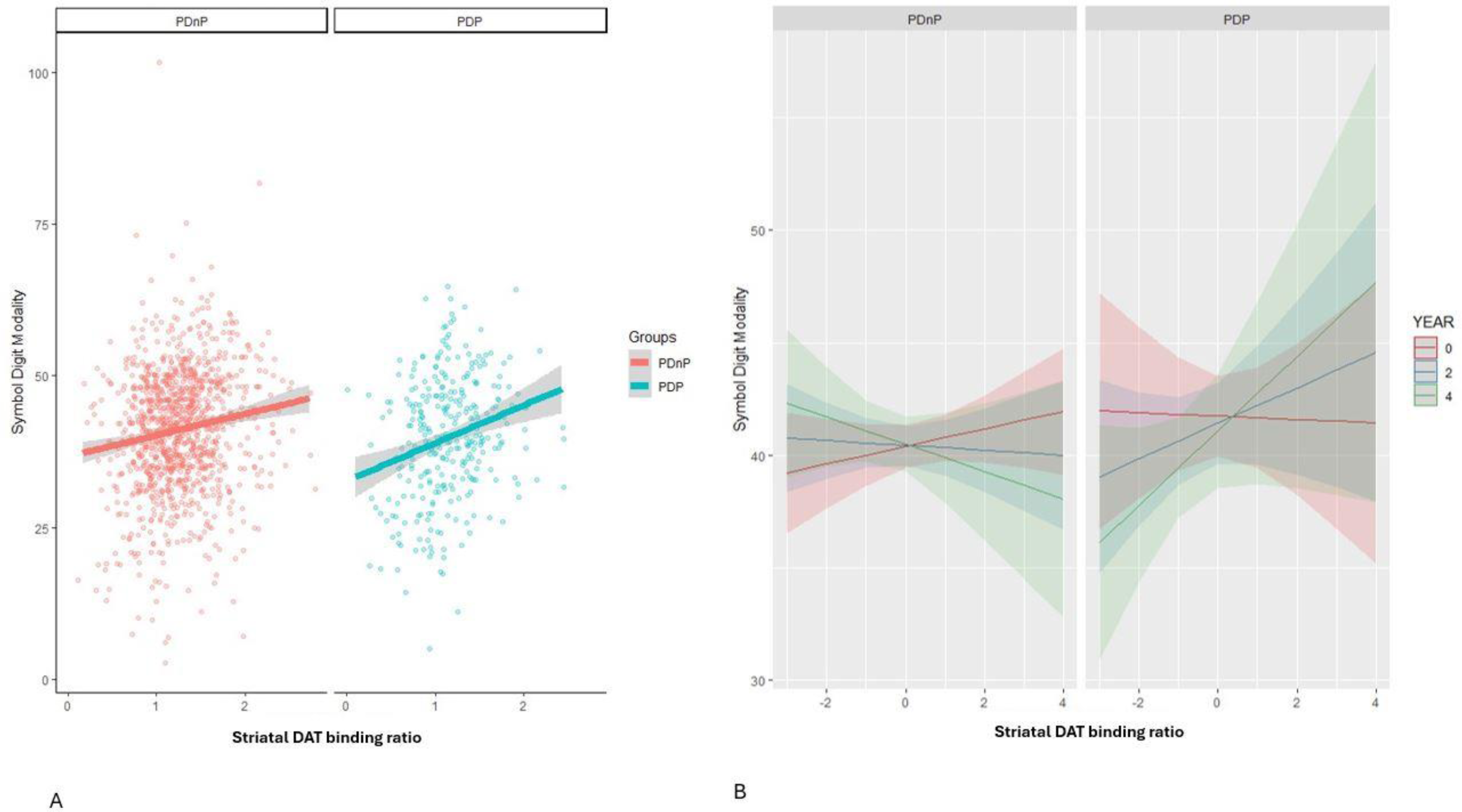
Trajectory of cognitive performance in PD psychosis in the Symbol Digit Modality across the years of the PPMI study and DAT binding ratio in striatum (A), and predictive values of SDM for each PD group (B).

## Discussion

Here, we examined whether the longitudinal trajectory of striatal presynaptic dopamine function as indexed by striatal DAT binding ratio was different between people with PD with and without psychosis and the extent to which this was associated with the longitudinal trajectory of cognitive performance in these two groups. As expected, striatal presynaptic dopamine function showed significant decline over time in both groups of PD. This is consistent with and extends previous reports ^15^ ^16^ to show that the same decremental pattern is evident even in PD patients who go on to develop psychosis. Further, here we show that PDP patients had significantly lower striatal presynaptic dopamine function compared to PDnP patients across all the yearly follow-up time-points over 4 years, extending previous evidence of lower striatal DAT binding from cross-sectional studies ^8–10^. However, contrary to our predictions, we did not find that the longitudinal trajectory of striatal DAT binding ratio differed between PD patients with and without psychosis. Finally, we found that accelerated longitudinal decline in Symbol Digit Modality performance in PDP patients compared to PDnP patients was differentially associated with the longitudinal trajectories of decline in striatal DAT binding ratio in the two groups even after controlling for the potential confounding effects of baseline sociodemographic characteristics and longitudinal course of depression, sleep changes, and motor symptom severity. We did not find a similar association between the accelerated longitudinal decline in MoCA performance in PDP compared with PDnP and longitudinal decline in striatal DAT binding ratio.

Our results may suggest that the differential longitudinal trajectories of Symbol Digit Modality and MoCA performance in PDP and PDnP may have different relationships to group trajectories of longitudinal decline in presynaptic dopamine function. More specifically, in people with PDnP, the nature of the relationship between Symbol Digit Modality performance and presynaptic dopamine function changed over time from being modestly but directly related in the early stages of PD to being inversely related at the later stages of illness. In contrast, in PD patients with psychosis the nature of the relationship changed in the opposite direction, such that from having minimal association in the early stages of the illness, there was a progressively stronger and direct correlation between Symbol Digit Modality performance and presynaptic dopamine function over the 4-year follow-up period. In contrast, the accelerated decline in MoCA in PDP compared to PDnP appeared to be unrelated to longitudinal decline in presynaptic dopamine function, as the longitudinal decline in MoCA scores remained significantly greater in PDP compared to PDnP even after controlling for the differential longitudinal trajectories of striatal DAT binding ratios in the two groups. While results presented here may suggest that differential decline in presynaptic dopamine function underlies the group difference (PDP vs PDnP) in longitudinal decline in Symbol Digit Modality, a cautious interpretation is warranted given the observational nature of the study. Further, we did not find a significant group difference in the longitudinal decline in presynaptic dopamine function, though one may speculate whether this reflects the relatively modest size of the cohort analysed here. The importance of sample size is also evident from our observation that group differences in the longitudinal trajectories of cognitive task performance was only significant for Symbol Digit Modality and MoCA in the present sample unlike in the larger sample from the same cohort wherein we also found an accelerated decline across the domains of visuo-spatial abilities, semantics aspects of language, immediate and delayed recall, in addition to MoCA and Symbol Digit Modality ^33^. As a result, we only tested the relationship between longitudinal trajectory of presynaptic dopamine function and longitudinal trajectory of MoCA and Symbol Digit Modality performance and not any of the other domains, which warrant testing in large samples.

Performance on the Symbol Digit Modality task relies on processing speed as well as associative learning and working memory ^50^ ^51^, and has been linked to activation in the frontoparietal attentional network, occipital cortex as well as cuneus, precuneus and cerebellum ^52^. In one conceptualisation ^53^, information processing speed combines sensory processing speed mediated in the primary visual cortex ^54^; cognitive speed engaging visuo-spatial attention mediated in the superior parietal lobule ^55^ ^56^, executive control and working memory mediated in Brodmann area 9 in the prefrontal cortex ^57^ and motor integration and attention mediated in the cerebellum ^58^; and finally motor speed as part of response preparation mediated in the Brodmann area 6 ^59^. In contrast, associative learning has been linked to medial temporal ^60^ as well as prefrontal, striatal and midbrain ^61–65^. Our result showing a differential relationship between the longitudinal trajectory of striatal presynaptic dopamine function and accelerated decline in symbol digit processing performance in people with PDP is consistent with evidence that neural substrates engaged during the Symbol Digit Modality task are implicated in the neurobiology of psychosis in PD ^1^ ^17^ ^66^ as well as evidence that presynaptic dopamine levels in the striatum may be linked with cognitive processes engaged by the Symbol Digit Modality task. Specifically, presynaptic dopamine levels in the caudate have been linked to working memory capacity and working memory load-related prefrontal activation, while levels in the putamen has been linked to motor speed in healthy individuals over the age of 60 ^67^. Independent evidence has linked also striatal dopamine function with processing speed ^68^ and attention, working memory, executive function and motor performance ^69^ ^70^ in people with PD. Further, phasic responses of brainstem dopaminergic nuclei have been shown to be involved in working memory updating and representation in the prefrontal cortex, suggesting a role for dopamine phasic signals in these processes ^62^.

What might underlie the differential association between striatal presynaptic dopamine function and accelerated decline in symbol digit processing performance in people with PD who develop psychosis? Given the early involvement of putamenal dopaminergic terminals in PD ^69^ and its role in motor speed, a cautious interpretation may be that as striatal presynaptic dopamine function deteriorates due to progressive degeneration of nigrostriatal neurons, people with PD who do not develop psychosis may increasingly rely on other strategies useful in maintaining processing speed by engaging regions involved in visuo-spatial attention, working memory and associative learning to continue processing new information. A speculative interpretation is that people with PD who develop psychosis and who have an even lower presynaptic dopamine function at baseline compared to PDnP, differ in their cognitive reserve ^71^ to engage these strategies and therefore experience a faster decline in processing speed despite not experiencing a faster decline in presynaptic striatal dopamine function compared to PDnP. It is worth noting that global cognition, as indexed by MoCA shows faster decline in PDP compared to PDnP even after accounting for the decline in striatal presynaptic dopamine function. Given the mixed evidence regarding benefits of cognitive training in people with PD and cognitive impairment ^72^, a common occurrence in people with PD psychosis ^73^, and the role of striatum in transfer of learning following training ^65^ our results underscore the importance of cognitive training strategies that engage cognitive processes with overlapping processing components and brain substrates.

## Strengths and limitations

Our work has some limitations. First of all, although the effect of dopamine-replacement medications on cognition is far from clear ^74–77^, we did not take into account PD medications, expressed as LEDD, in our analyses as the PD groups did not differ on the amount of LEDD taken across the 4 years of the PPMI study (see Supplementary Material 1). Nevertheless, we cannot be certain that exposure to dopamine agonists may have confounded the results presented here. It may also be argued that our approach to categorisation of people with PD psychosis ^48^ may not be as sensitive as classification on the basis of measures such as the Schedule for Assessment of Positive Symptoms in PD (SAPS-PD ^78^) which allows for more specific grouping, such as patients with hallucinations and/or with delusions. It is worth noting that we did not test the relationships between trajectory of presynaptic dopamine function and performance in other cognitive tasks, specifically those providing a more direct measure of working memory and executive function, for reasons explained before. Results presented here underscore the importance of testing these relationships as well, in larger samples. Another limitation worth considering is related to the limitation of the PPMI cohort in terms of lack of diversity with regard to ethnic origin, being composed predominantly of individuals from a White ethnic background ^79^ ^80^. It has been reported that PD patients from a Black ethnic background are more likely to experience cognitive deficits and later develop dementia compared to other ethnic minorities ^81^ ^82^, whilst the evidence for PD patients from Asian background remains ambiguous ^83^. Whether ethnic background may also impact the longitudinal trajectory of striatal DAT uptake or its relationship with cognitive decline in PD patients with psychotic symptoms remains to be tested. Finally, while we have focused on a proxy index of presynaptic striatal dopamine function, we cannot rule out the role of other neurotransmitters underlying these longitudinal changes in cognition ^17^, which warrant investigation in future studies.

## Conclusion

In conclusion, our results suggest that longitudinal changes in striatal presynaptic dopamine function may underlie the greater longitudinal decline in performance in the symbol digit modality task, that engages processing speed, associative learning and working memory processes, but not a similar differential longitudinal course of decline in general cognitive ability in people with PD who develop psychosis compared to those who do not PDP and PDnP. Whether striatal presynaptic dopamine changes explain accelerated longitudinal decline in other cognitive domains in people with PDP remains to be tested.

## CRediT authors’ contributions

*Conceptualisation:* Sara Pisani, Sagnik Bhattacharyya

*Methodology:* Sara Pisani, Sagnik Bhattacharyya

*Investigation:* Sara Pisani, Latha Velayudhan, Dominic ffytche, Sagnik Bhattacharyya

*Data curation:* Sara Pisani, Sagnik Bhattacharyya *Formal analysis:* Sara Pisani, Sagnik Bhattacharyya *Visualisation*: Sara Pisani, Sagnik Bhattacharyya

*Funding acquisition*: Sagnik Bhattacharyya, Latha Velayudhan, Dominic ffytche, Dag Aarsland, Kallol Ray Chaudhuri, Clive Ballard

*Writing (original draft):* Sara Pisani, Latha Velayudhan, Sagnik Bhattacharyya

*Writing (review & editing):* All authors

*Supervision*: Latha Velayudhan, Dominic ffytche, Sagnik Bhattacharyya

## Data Availability

Data used in the preparation of this article were obtained [on February 1st 2023] from the Parkinson's Progression Markers Initiative (PPMI) database (www.ppmi-info.org/access-dataspecimens/download-data), RRID:SCR 006431. For up-to-date information on the study, visit www.ppmi-info.org.

## Acknowledgement

Data used in this work were obtained from the Parkinson’s Progression Markers Initiative database (http://www.ppmi-info.org/). PPMI – a public-private partnership – is funded by the Michael J. Fox Foundation for Parkinson’s Research and funding partners, including 4D Pharma, Abbvie, AcureX, Allergan, Amathus Therapeutics, Aligning Science Across Parkinson’s, AskBio, Avid Radiopharmaceuticals, BIAL, Biogen, Biohaven, BioLegend, BlueRock Therapeutics, Bristol-Myers Squibb, Calico Labs, Celgene, Cerevel Therapeutics, Coave Therapeutics, DaCapo Brainscience, Denali, Edmond J. Safra Foundation, Eli Lilly, Gain Therapeutics, GE HealthCare, Genentech, GSK, Golub Capital, Handl Therapeutics, Insitro, Janssen Neuroscience, Lundbeck, Merck, Meso Scale Discovery, Mission Therapeutics, Neurocrine Biosciences, Pfizer, Piramal, Prevail Therapeutics, Roche, Sanofi, Servier, Sun Pharma Advanced Research Company, Takeda, Teva, UCB, Vanqua Bio, Verily, Voyager Therapeutics, the Weston Family Foundation and Yumanity Therapeutics. For up-to-date information on the study, visit www.ppmi-info.org.

## Data availability statement

Data used in the preparation of this article were obtained [on February 1^st^ 2023] from the Parkinson’s Progression Markers Initiative (PPMI) database (www.ppmi-info.org/access-dataspecimens/download-data), RRID:SCR 006431. For up-to-date information on the study, visit www.ppmi-info.org.

