## Supplementary Material for "Decline in striatal binding ratio associated with accelerated decline in performance on Symbol Digit Modality but not MoCA in Parkinson’s disease psychosis"

### Supplementary Material 1

#### *Levodopa equivalent daily dose (LEDD)*

PD medications were recorded across all study visits. Data on dopamine-replacement medication, expressed as Levodopa equivalent daily dose (LEDD). The PPMI study team has reported in a detailed document the different conversion factors, and the procedures. Please visit their website www.ppmi-info.org/study-design for more information. eTable1 shows of the amount of LEDD for each patient's group from year 1 to year 4 follow up. eTable2 reports the number of patients per study year, and eTable3 reports the number (and percentage) of people who reported psychotic symptoms per study year.

There were no overall differences between PDnP and PDP ($p$=0.990), or between PDnP and PDP at baseline ($p$=0.873) in LEDD. LEDD amount increased significantly across the years, on similar degree for all three patients' groups (t=12.872, b=110.841, $p$<0.001).

**eTable1.** Means and SDs of LEDD for PD patients (PDnP, PDP and PDP at baseline).

| Study year | Group (n, number of patients) | LEDD (mean ± SD) |
|---|---|---|
| Year 1 | PDnP (n=295) | 318.265± 251.397 |
| | PDP (n=82) | 240.271 ± 152.292 |
| | PDP at baseline (n=9) | 250 ± 150 |
| Year 2 | PDnP (n=287) | 387.802 ± 312.597 |
| | PDP (n=79) | 398.258 ± 294.299 |
| | PDP at baseline (n=6) | 312.5 ± 17.678 |
| Year 3 | PDnP (n=272) | 480.422 ± 463.857 |
| | PDP (n=77) | 521.659 ± 348.087 |
| | PDP at baseline (n=9) | 485.667 ± 354.785 |
| Year 4 | PDnP (n=252) | 563.626 ± 471.843 |
| | PDP (n=73) | 571.438 ± 264.884 |
| | PDP at baseline (n=7) | 422.8 ± 374.432 |

#### *Differences between PD groups*

**eTable2.** Number of patients per study year from baseline to year 4 (PDnP, PDP and PDP at baseline).

| | Baseline | Year 1 | Year 2 | Year 3 | Year 4 |
|---|---|---|---|---|---|
| PD patients without psychosis (PDnP) | 325 | 295 | 287 | 272 | 252 |
| PD patients with psychosis (PDP) | 83 | 82 | 79 | 77 | 73 |
| PD patients with psychosis from baseline (PDP at baseline) | 12 | 9 | 6 | 9 | 7 |

**eTable 3.** Number and percentage of patients who reported psychotic symptoms over the years of the PPMI study.

| | PD patients | PD patients who reported psychotic symptoms (n, %) |
|---|---|---|
| Baseline | 420 | 12 (2.86%) |
| Year 1 | 386 | 16 (4.15%) |
| Year 2 | 372 | 15 (4.03%) |
| Year 3 | 358 | 21 (5.87%) |
| Year 4 | 332 | 15 (4.52%) |

**eTable 4.** Sample baseline characteristics of the three groups, i.e., PD patients without psychosis (PDnP), PD patients with psychosis (PDP), and PD patients who developed psychotic symptoms at baseline (PDP at baseline).

| | PDnP (n=325) | PDP (n=83) | PDP at baseline (n=12) | Significance test * |
|---|---|---|---|---|
| Age | 61.550 ± 10.022 | 62.443 ± 8.477 | 58.405 ± 9.364 | F(2)=0.962, p=0.383 |
| Sex (n, %) | 214 (65.8%) | 51 (61.4%) | 10 (83.3%) | χ2 (2)=2.309, p=0.315 |
| Ethnicity (n, %) | | | | χ2 (6)=10.657, p=0.093 |
| *White (n, %)* | 304 (93.5%) | 73 (87.9%) | 11 (91.7%) | |
| *Black (n, %)* | 2 (0.6%) | 4 (4.8%) | 0 | |
| *Asian (n, %)* | 5 (1.5%) | 3 (3.6%) | 0 | |
| *Other (n, %)* | 14 (4.3%) | 3 (3.6%) | 1 (8.3%) | |
| PD onset (age in years) | 59.648 ± 10.140 | 60.220 ± 8.692 | 54.850 ± 12.953 | F(2)=1.528, p=0.218 |
| PD diagnosis (age in years) | 61.003 ± 9.991 | 61.876 ± 8.452 | 57.735 ± 9.471 | F(2)=1.005, p=0.367 |
| Years of education | 15.686 ± 3.013 | 15.084 ± 2.859 | 15.833 ± 2.038 | F(2)=0.043, p=0.958 |
| PD duration (months) | 6.574 ± 6.300 | 6.814 ± 6.955 | 8.031 ± 9.208 | F(2)=0.316, p=0.729 |
| MoCA | 27.080 ± 2.285 | 27.446 ± 2.318 | 26.333 ± 2.964 | F(2)=1.563, p=0.211 |
| HVLT immediate recall | 24.454 ± 4.906 | 24.349 ± 5.269 | 24.083 ± 5.485 | F(2)=0.043, p=0.958 |
| HVLT delayed recall | 8.324 ± 2.550 | 8.410 ± 2.518 | 8.917 ± 1.881 | F(2)=0.339, p=0.712 |
| HVLT discrimination | 9.712 ± 2.610 | 9.410 ± 2.687 | 9.250 ± 2.927 | F(2)=0.57, p=0.566 |
| HVLT recognition | 11.161 ± 1.295 | 11.241 ± 1.031 | 11.167 ± 0.718 | F(2)=0.139, p=0.87 |
| Letter number sequence | 10.639 ± 2.755 | 10.289 ± 2.081 | 11.250 ± 3.571 | χ2 (2)=1.038, p=0.595 |
| Benton judgement of line orientation (BJLOT) | 12.855 ± 2.135 | 12.530 ± 2.132 | 12.500 ± 1.784 | F(20)=0.879, p=0.416 |
| Symbol digit modality (SDM) | 41.222 ± 9.596 | 41.169 ± 9.146 | 42.167 ± 16.022 | χ2 (2)=0.020, p=0.989 |
| Semantic fluency test (total) | 48.386 ± 11.409 | 50.193 ± 12.741 | 46.583 ± 10.858 | F(2)=0.994, p=0.371 |
| Depression (GDS) | **2.142 ± 2.312** | 2.831 ± 2.709 | **3.833 ± 3.353** | **F(2)=5.051, p=0.007** [b] |
| Sleep (ESS) | 5.520 ± 3.214 | 6.313 ± 3.803 | 8.667 ± 5.051 | χ2 (2)=6.343, p=0.042 |
| REM behaviour | **3.882 ± 2.556** | **4.687 ± 2.789** | **6.500 ± 3.555** | **F(2)=8.129, p<0.001** [a,b] |
| Anxiety (STAI) | **64.031 ± 17.672** | 68.301 ± 19.650 | **80.333 ± 19.942** | **F(2)=6.048, p=0.003** [b] |
| UPDRS part I scores | **5.108 ± 3.830** | **6.807 ± 4.326** | **9.333 ± 5.245** | **F(2)=11.58, p<0.001** [a,b] |

| | | | | |
|---|---|---|---|---|
| UPDRS part II scores | **5.565 ± 3.986** | **6.952 ± 4.504** | **6.000 ± 5.187** | **F(2)=3.736, p=0.025** [a] |
| UPDRS part III scores | 20.465 ± 8.755 | 22.108 ± 8.803 | 23.667 ± 11.949 | F(2)=1.747, p=0.176 |
| Rigidity ** | 3.603 ± 2.555 | 4.253 ± 2.749 | 4.917 ± 3.704 | χ2 (2)=5.424, p=0.066 |
| SCOPA-Autonomic | **8.630 ± 5.522** | **12.444 ± 7.136** | 13.000 ± 8.954 | **χ2 (2)=22.95, p<0.001** [a] |
| Tremor *** | 4.308 ± 2.994 | 4.578 ± 3.778 | 4.583 ± 2.234 | F(2)=0.273, p=0.761 |

a PDnP vs PDP
b PDnP vs PDP at baseline
c PDP vs PDP at baseline

*When applicable, we applied the non-parametric equivalent of a one-way analysis of variance, i.e., Kruskal-Wallis (χ2)
** Rigidity scores were derived from the MDS-UPDRS part III. They represent the total score of rigidity item for neck, right and left upper extremities, and right and left lower extremities.
*** Tremor scores were derived from the MDS-UPDRS part III. They represent the total score of "postural tremor of hands (right and left)", "kinetic tremor of hands (right and left)", "rest tremor amplitude (right upper extremities, left upper extremities, right lower extremities, left lower extremities, lip/jaw)", and "consistency of tremor".
